# Predictive value of Acute Physiology and Chronic Health Evaluation II (APACHE II) scoring in patients admitted to intensive care units

**DOI:** 10.64898/2025.12.31.25343298

**Authors:** Saurav Jha, Shirjan Gautam, Samir Shiwakoti

**Affiliations:** Department of Internal Medicine, Kathmandu Medical College, Kathmandu, Bagmati Province, Nepal; Department of General Surgery, Kathmandu Medical College, Kathmandu, Bagmati Province, Nepal

## Abstract

**Background:** Risk stratification tools are essential for guiding care and allocating limited resources in intensive care units (ICUs). Evidence on the performance of the Acute Physiology and Chronic Health Evaluation II (APACHE II) score in Nepal is limited. We evaluated the association of APACHE II measured within 24 hours of ICU admission with mortality, ICU length of stay, and discharge disposition in a tertiary hospital in Nepal.

**Methods and findings:** We conducted a prospective observational cohort study of consecutive adult patients admitted to three multidisciplinary ICUs at a tertiary hospital in Kathmandu, Nepal. Patients with ICU stay <24 hours and those admitted to coronary or neonatal ICUs were excluded. APACHE II was calculated from the worst values in the first 24 hours. The primary outcome was ICU mortality; secondary outcomes were ICU length of stay and discharge disposition (home, ward, high-care/step-down, or in-hospital death). Discrimination for mortality was assessed using receiver operating characteristic (ROC) analysis. Associations with length of stay were examined using linear regression; discharge disposition was evaluated using multinomial logistic regression.

Among 200 patients (54% male; mean age 54.7 ± 21.0 years), ICU mortality was 23.0% (46/200). Non-survivors were older than survivors (mean difference 13.31 years; 95% CI 6.60–20.02; p < 0.001). The mean APACHE II score was 13.19 ± 7.89 overall, higher in non-survivors vs survivors (19.85 ± 7.14 vs 11.21 ± 6.98; p < 0.001). APACHE II discriminated ICU mortality well (AUC 0.806); a cutoff ≥14.5 yielded 76.1% sensitivity and 71.4% specificity. Mortality increased across APACHE II strata (trend p < 0.001). Glasgow Coma Scale scores were lower in non-survivors (10.85 ± 4.43) than survivors (13.29 ± 3.11; p < 0.001). Higher APACHE II scores were associated with in-hospital death versus ward discharge, but not with high-care versus ward (OR 1.063; 95% CI 0.994–1.136; p = 0.074). Limitations include the single-center design, modest sample size, and short study duration, which may limit generalizability.

**Conclusions:** APACHE II measured within 24 hours of ICU admission demonstrated good discrimination for ICU mortality and a modest association with ICU length of stay in a Nepalese tertiary setting, while being less informative for intermediate discharge dispositions. Together with Glasgow Coma Scale, APACHE II offers a pragmatic, cost-effective approach to early prognostication and resource planning in resource-limited ICUs. Multicenter studies are warranted to validate these findings across Nepal.

## Introduction

According to Painter et al, “A critically ill or injured patient is defined as one who has an illness or injury impairing one or more vital organ systems such that there is a high probability of imminent or life-threatening deterioration in the patient’s condition.” [1] It is important to stratify critically ill patients into risk groups to guide their admission into respective units in the hospital and the treatment they are going to receive.[2] Working in an intensive care unit (ICU) requires making early predictions about patient outcomes within the first 24 hours of admission. This involves estimating the likelihood of mortality by analyzing key variables typically used in diagnosing and treating critically ill patients.[3] Some widely used scoring systems for critically ill ICU patients are the Acute Physiology and Chronic Health Evaluation (APACHE) I-IV, the Mortality Prediction Model (MPM) and the Simplified Acute Physiology Score (SAPS) I-III.[4,5] APACHE scoring system was first introduced in 1981 AD by William Knaus et al and was updated 4 years later to APACHE II.[5–7] The APACHE-II system assigns points for 12 physiologic variables, for age, and for chronic health status, in generating a total point score. While measuring APACHE II, patient’s temperature, heart rate, respiratory rate, mean arterial blood pressure, oxygenation, arterial pH, serum potassium, sodium, and creatinine, hematocrit, white blood cell (WBC), and Glasgow Coma Scale are measured.[8] These 12 variables are used to assign an integer score from 0 to 71 for age, and for chronic health status. [9,10]

The APACHE scoring system was developed in USA and its application may or may not be equally applicable throughout different countries of the globe, especially in the context of Nepal where the literature regarding the study of APACHE II for predicting mortality in ICU settings is limited.[11] This study was conducted to study the correlation between the APACHE II score within 24 hours of admission into ICU and length of ICU stay, outcome of mortality and discharge disposition.

## Methods

We conducted a prospective observational cohort study in Kathmandu Medical College and Teaching hospital, a tertiary care center in Kathmandu, Nepal. There are a total of four ICUs in the hospital, including a neonatal ICU. Data were taken from three ICU, and neonatal ICU was excluded. The ICUs were mixed medical-surgical units providing level III critical care. Patients with missing data required to calculate APACHE II score were excluded as well. Ethical Approval was obtained from the Institutional Review Committee of the hospital with reference no. 21072025/26. This research was conducted in accordance with the Declaration of Helsinki. The requirement for written informed consent was waived by the institutional review committee as only de-identified patient data were collected. Patients were admitted to these ICUs based on whether they had existing or impending organ failure, or if they underwent a recent surgery at our center. The ICUs were handled by dedicated critical care experts, including critical care residents, consultants from different faculty throughout the hospital, on-call anesthesia residents, junior residents, medical officers, and trained nurses.

We included patients admitted to the ICU and excluded patients admitted to coronary care units, and neonatal ICUs. The sample size was calculated to estimate the diagnostic performance of APACHE II score for predicting ICU mortality. Published metanalyses were used to derive the estimates. A pooled sensitivity of 88%, specificity of 84%, and an AUC of 0.84 for APACHE II in predicting mortality was used.[12] An expected ICU mortality prevalence of 29.6% was used.[13] Based on these parameters, the sample size required to estimate sensitivity with 95% confidence and ±10% precision was 138 patients, which exceeded the number required for estimating specificity or ROC-AUC. To account for approximately 10% incomplete data, the target sample size was increased to 152 patients. However, the study included 200 patients to increase the precision of the analysis and reduce random variability. Bias was minimized using validated scoring systems, structured data collection, consecutive patients inclusion, and stratification. Recall and misclassification bias were also limited as all information was extracted from clinical records. Multiple approaches were used to identify and minimize the potential confounders within the data set. These approaches included comparing baseline characteristics between survivors and non-survivors, assessing correlations between key variables such as age and APACHE II score, examining outcome patterns through stratified APACHE II categories and regression analyses.

A prospective study was carried out in our hospital from 1^st^ August 2025 to 30^th^ November 2025. The data of the patients admitted into different intensive care units of our hospital were recorded over the period of 4 months. Data were recorded on a daily basis within 24 hours of admission. Patients whose ICU stay was less than 24 hours were excluded from the study. The variables included were the age, sex and ethnicity of the patient, the date of admission, the primary diagnosis, any co-morbidities that the patient had, their mean arterial pressure, body temperature, arterial pH, heart rate, respiratory rate, serum sodium, potassium, creatinine, hematocrit, WBC count, GCS score, FiO2, PaO2, PaCO2 and eventually their calculated APACHE II score. The length of ICU stay and the outcome, i.e. mortality or discharge disposition was also recorded. The primary outcome was ICU mortality; secondary outcomes were ICU length of stay and discharge disposition (home, ward, high-care/step-down, or in-hospital death). ICU mortality was defined as death occurring during the ICU stay prior to discharge or transfer. High-care units were defined as step down units providing continuous monitoring without invasive organ support.

The data of the aforementioned variables were entered into SPSS version 26 and the results were extrapolated and are mentioned in this study.

## Result

A total of 200 patients were included in this study. 108 (54%) of them were men and 92 (46%) were females. The mean age of participants was 54.7 ± 21 years (Range 19-98). 46 (23%) of the patients died while 154 (77%) of patients survived. There was a statistically significant difference in the mean age of survivors (51.63 ± 20.68 years) as compared to non-survivors (64.93 ± 18.73 years). The mean age difference was 13.31 years (95% CI: 6.60 to 20.02; *p* < 0.001).

The mean APACHE II score of all patients admitted in the ICU was 13.19 ± 7.89. The mean APACHE II score among non-survivors was 19.85 ± 7.14 and found to be significantly higher (p<0.001) than survivors with score of 11.21 ± 6.98. A moderate positive correlation between age and APACHE II score was found which was statistically significant (r = 0.540, p < 0.001).

The predictive value of APACHE II scoring system was evaluated using area under ROC curve. It was found that the APACHE II score significantly predicted ICU mortality with an area under curve of 0.806 which indicated good diagnostic accuracy, as shown in Fig 1. The optimal value for cut off score for predicting mortality was ≥14.5 with sensitivity of 76.1% and specificity of 71.4%.

**Fig 1.**
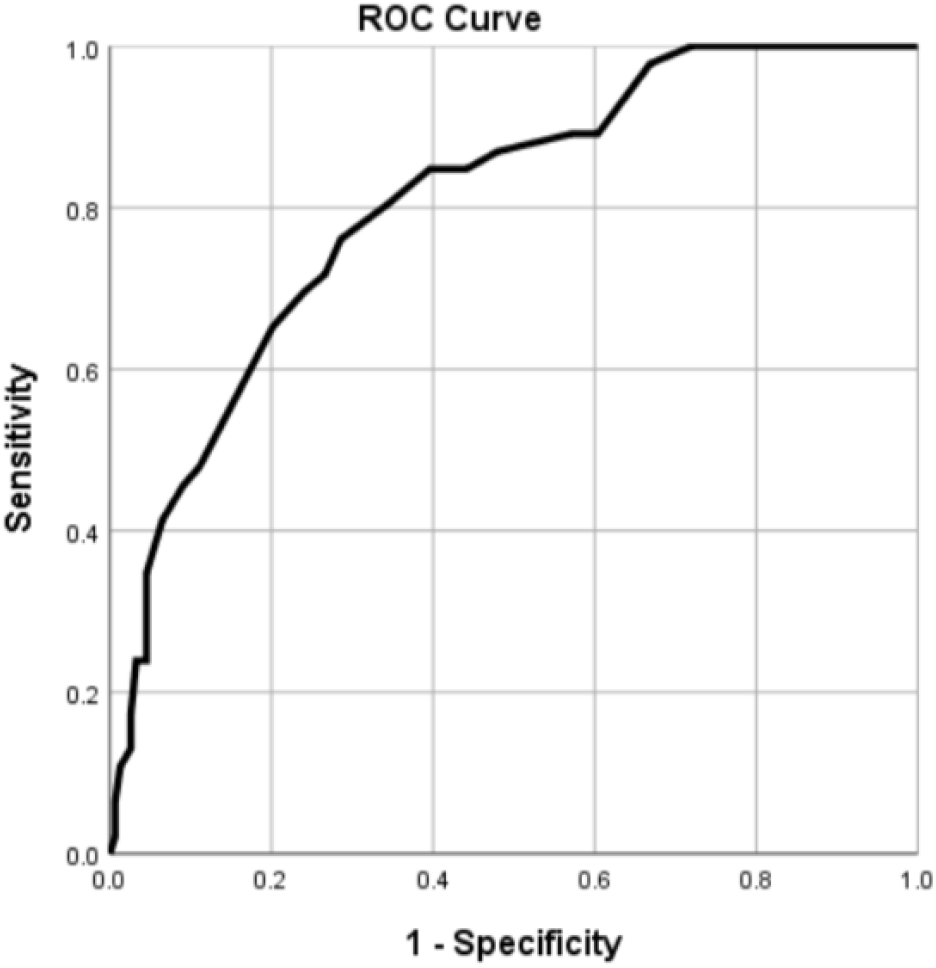
Receiver Operating Characteristic (ROC) Curve for APACHE II Score in Predicting ICU Mortality.

The mortality rates according to APACHE II Scores of the patients are shown in the Table 1. Chi-square test showed statistically significant association between APACHE II score groups and mortality (χ^2^= 42.723, *p* < .001) pointing out that there was significant difference in the mortality rates across APACHE II score groups. The linear-by-linear association test was also significant (χ^2^ = 39.218, *p* < .001), suggesting a trend of increasing mortality with higher APACHE II scores.

**Table 1.**
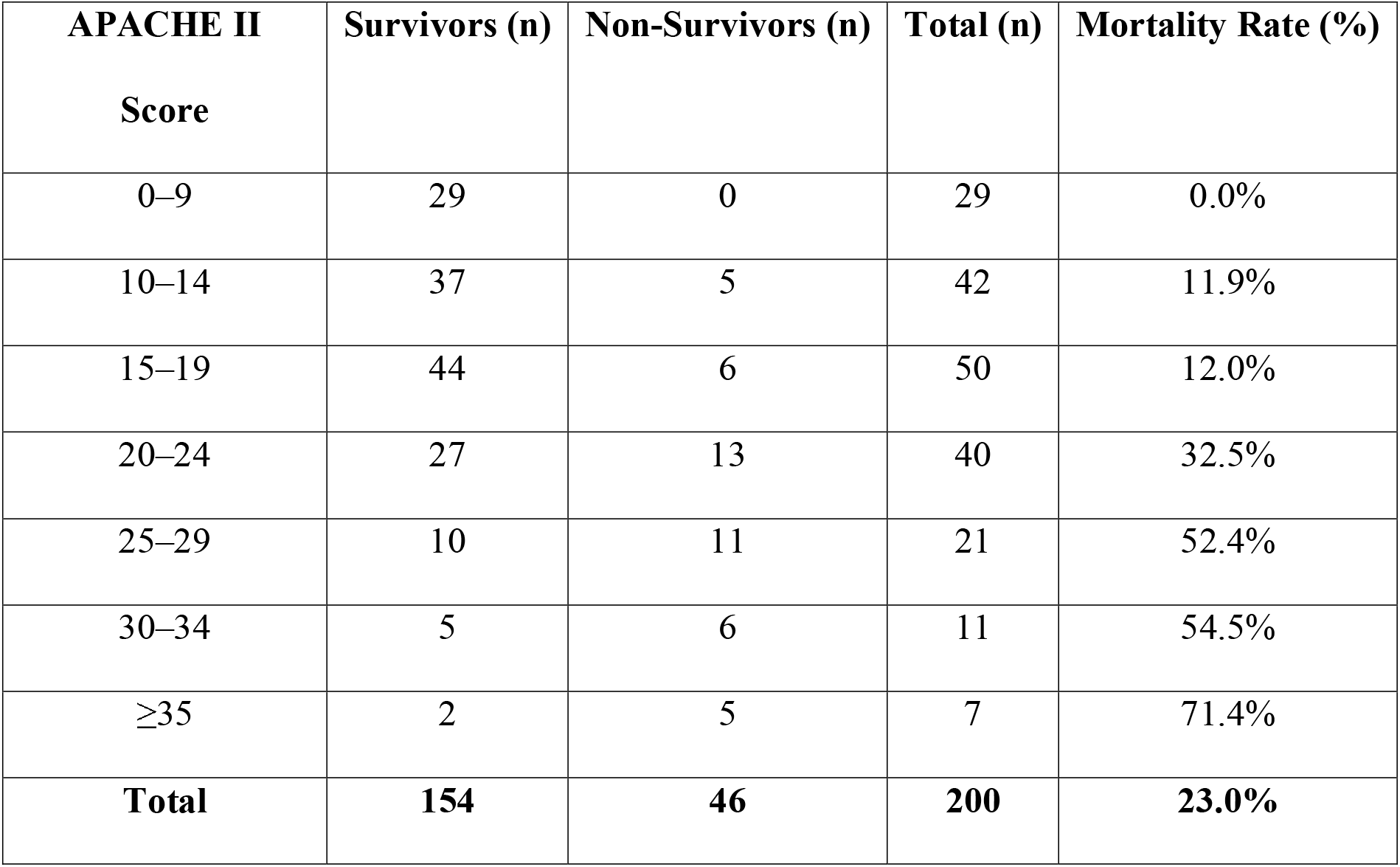
Distribution of ICU Mortality Across APACHE II Score Categories.

The mean Glasgow Coma Scale (GCS) score among survivors was 13.29 ± 3.11, while among non-survivors it was 10.85 ± 4.43 and the difference was statistically significant (p<0.001). Linear regression showed that APACHE II score was a significant predictor of ICU stay duration (B= 0.153, p<0.001). A weak positive correlation between APACHE II score and ICU stay was observed (R=0.298). About 9% of variance in the ICU stay was explained by the model.

A multinomial logistic regression was performed to evaluate whether APACHE II score can predict discharge disposition to High-care, Ward or Home. The model was statistically significant, χ^2^(2) = 47.61, *p* < .001. Higher APACHE II scores were significantly associated with in-hospital deaths compared to patients discharged to the ward. However, there was no statistically significant association between APACHE II score and discharge disposition to high-care unit vs. Ward. (OR = 1.063, 95% CI :0.994 to 1.136, p = 0.074).

The APACHE II Scores and Mortality rate according to the disease category of the patients is shown in Table 2. The patients with respiratory causes had the highest mortality rate of 35.5% with APACHE II score ranging 16.55 ± 7.02 closely followed by cardiovascular causes with mortality of 33.3% and APACHE II score of 16.00 ± 8.86.

**Table 2.** APACHE II Score and Mortality rate among different disease categories.

| Disease Category | APACHE II Score (Mean ± SD) | Mortality Rate (%) |
| --- | --- | --- |
| Neurological Causes | $11.03 \pm 6.29$ | 18.2% |
| Respiratory Causes | $16.55 \pm 7.02$ | 35.5% |
| Gastrointestinal Causes | $10.67 \pm 6.98$ | 14.3% |
| Cardiovascular Causes | $16.00 \pm 8.86$ | 33.3% |
| Sepsis/Septic Shock | $13.38 \pm 5.10$ | 25.0% |
| Other | $11.35 \pm 9.42$ | 12.5% |
| **Total** | **$13.20 \pm 7.89$** | **23.0%** |

## Discussion

The mean age of survivors was significantly lower than non-survivors. However, the positive correlation between APACHE II score and age suggests that aging is associated with increased vulnerability to critical illness. This finding was similar to other studies which imply APACHE II score is the predictor of mortality and not the age.[14,15] In countries like Nepal, where access to health care among elderly may be limited, the impact of age on disease severity and outcome may be even more detrimental.[16]

Our study concluded that APACHE II score significantly predicted ICU mortality which is also supported by many similar studies.[17–19] In our study, the cut off value of ≥14.5 was found to predict ICU mortality with a sensitivity of 76.1% and specificity of 71.4%, demonstrating balanced and good predictive accuracy. A wide range of studies have shown APACHE II score having sensitivity from 51% to 93% and specificity from 49% to 97%.[20] Our findings are parallel to the findings by Yan et al. (2021) in a study among gram-negative sepsis patients with APACHE II cut off of 18.5 with sensitivity of 65.7% and specificity of 72.7%. [21]

In our study, stratified analysis using categorized APACHE II scores showed a clear trend of increasing ICU mortality with higher APACHE II scores. Mortality rose from 0% in lowest score group (0-9) to 71.4% in the highest score group (≥35). These findings are consistent with other researches indicating that APACHE II score is a reliable predictor of mortality in intensive care settings.[22,23.]For instance, a cohort study in Pakistan found that all the patients with APACHE II score ≥21 died, while around 70% of survivors had score of 11-20.[24] This weighs in on a clear stratification of outcomes based on score categories. These results support the idea of integrating APACHE II into ICU triage for risk assessment, guiding clinical decisions, and prioritizing resource allocation in resource-limited settings.

Survivors had a significantly higher mean GCS compared to non-survivors, indicating that lower GCS on admission is associated with higher mortality. This is in alignment with the findings of a meta-analysis of Ethiopian ICUs where patients with GCS below 9 were 4.7 times more likely to die in the ICU. [25] The

Pan-Asia Outcomes Study, evaluating over 16000 patients concluded that both full GCS and motor-only component of GCS has outstanding predictive accuracy for a 30-day mortality.[26] In resource-limited Nepalese settings, where advanced diagnostic tools may not be readily available, GCS can be a defining tool for early risk assessment and management.

The APACHE II score was a statistically significant, though modest, predictor of ICU stay duration. While it accounts for a small portion of the variance, higher scores were associated with slightly longer ICU stays. This supports the utility of the APACHE II score not only in predicting mortality but also in anticipating resource utilization in terms of ICU days. In contrast, a study conducted in Pakistan demonstrated an inverse, although weak, correlation between APACHE II and ICU stay pointing out that sicker patients often have shorter stays due to higher mortality. [27] Similarly a Japanese study concluded that increasing APACHE II scores were significantly associated with prolonged ICU stays.[28] These widely different results point to the fact that the relationship of APACHE II with length of ICU stay may be context-dependent. In some cases, higher scores mean greater severity and longer need of ICU care, while in others they may indicate terminal illness and shorter ICU stays due to increasing mortality. In our Nepalese ICU context, the positive association likely reflects a system where higher-severity illness corresponds to longer ICU resource utilization but is still tempered by an overall high mortality.

In this study, highest mortality of 35.5% was seen in patients with respiratory diseases closely followed by patients with cardiovascular diseases with a 33.3% mortality which is similar to the findings of Evran et. al with 30.7% and 16.5% mortality in patients with respiratory and cardiovascular diseases respectively being the first and second highest.[29] Sepsis had a mortality rate of 25% with lower mean APACHE II score which is in accordance with the global averages of 20-40%.[30]

## Limitations

The major limitation of this study is that it is a single center study so its outcomes may not be generalized to all the ICUs of Nepal. This study also has a small sample size which may limit its statistical power. Its short study duration may not be able to capture seasonal variations in ICU admissions. Comparison with other severity scores like SOFA or SAPS was not performed.

## Conclusion

Our study concludes that APACHE II score is a significant predictor of ICU mortality, modest predictor of length of ICU stay and less predictive of intermediate discharge outcomes. Higher APACHE II scores lead to higher mortality with a cut-off value of ≥ 14.5 demonstrating good diagnostic accuracy. Moreover, lower GCS scores were significantly associated with mortality. APACHE II and GCS can be used as a practical and cost-effective tool for early prognosis assessment and resource allocation in the ICUs of Nepal where resources and advanced health care is limited. Further multicenter studies with larger sample size are needed to validate and extend the findings of this research.

## Data Availability

The data supporting the findings of this study are available from the corresponding author upon reasonable request.

